# Initial development and pragmatic clinical validation of a static disease severity instrument for pyoderma gangrenosum: Investigator Global Assessment for PG (IGAPg)

**DOI:** 10.64898/2025.12.26.25342857

**Authors:** Michael E. Jacobson, Justin W. Ng, Laura Morales Leon, Jesse J. Keller, Angelo V. Marzano, William W. Huang, Robert I Kelly, Arash Mostaghimi, Alex G. Ortega-Loayza

## Abstract

**Background:** Pyoderma gangrenosum (PG) is a rare neutrophilic ulcerative dermatosis with no FDA-approved therapies and limited validated outcome measures. Investigator’s Global Assessments (IGAs) are widely used in dermatology but often lack objective criteria, robust validation, and comparability across studies. There remains a critical need for a PG-specific, standardized, and validated severity instrument.

**Methods:** We developed and conducted initial validation of the Investigator Global Assessment for Pyoderma Gangrenosum (IGAPg©), a novel PG-specific IGA. An international multistakeholder panel guided development, with a core team designing the instrument based on prior research identifying key objective clinical features of PG severity. The IGAPg© incorporates ulcer depth, drainage, discoloration, and undermining, with ulcer location and extent used to resolve indeterminate cases. Standardized rater training materials were created. Construct validity was assessed in 36 patients evaluated by an expert PG clinician, with correlations to Patient Global Assessment (PGA), Skindex-Mini, and numeric rating scales (NRS) for 24-hour and 7-day pain. Inter-rater reliability was evaluated in a subset of 26 patients assessed independently by five raters using a two-way random-effects intraclass correlation coefficient (ICC [2,1]) within a linear mixed-effects model.

**Result:** IGAPg© scores demonstrated strong correlation with PGA when assessed by an expert PG dermatologist (Pearson’s r = 0.73) and when averaged across all raters (r = 0.69). Moderate correlations were observed with Skindex-Mini (r = 0.49), 24-hour pain NRS (r = 0.48), and 7-day pain NRS (r = 0.52), consistent with differing constructs measured. Inter-rater reliability was high (ICC = 0.76). Raters reported the instrument to be comprehensive, comprehensible, and efficient to administer. Reliability and construct validity metrics are summarized in Table 1.

**Conclusions:** Despite limitations including modest sample size and rater homogeneity, the IGAPg© demonstrated strong construct validity and high inter-rater reliability. Designed for dermatologists and trainees familiar with PG morphology, the IGAPg© represents a promising PG-specific outcome measure for clinical research and therapeutic trials. Future work will focus on refining training materials and expanding validation with structured patient–investigator engagement.

## Dear Editor

Pyoderma Gangrenosum (PG) is a rare neutrophilic ulcerative dermatosis with no FDA-approved therapies and limited validated outcome measures.^1^ Investigator’s Global Assessments (IGAs) are widely used but often lack objective criteria and robust validation. ^2,3^ The use of multiple IGAs for a single condition further limits comparability. ^2^ Newer IGAs with objective criteria are being developed across dermatology.^3^

**Table 1.**
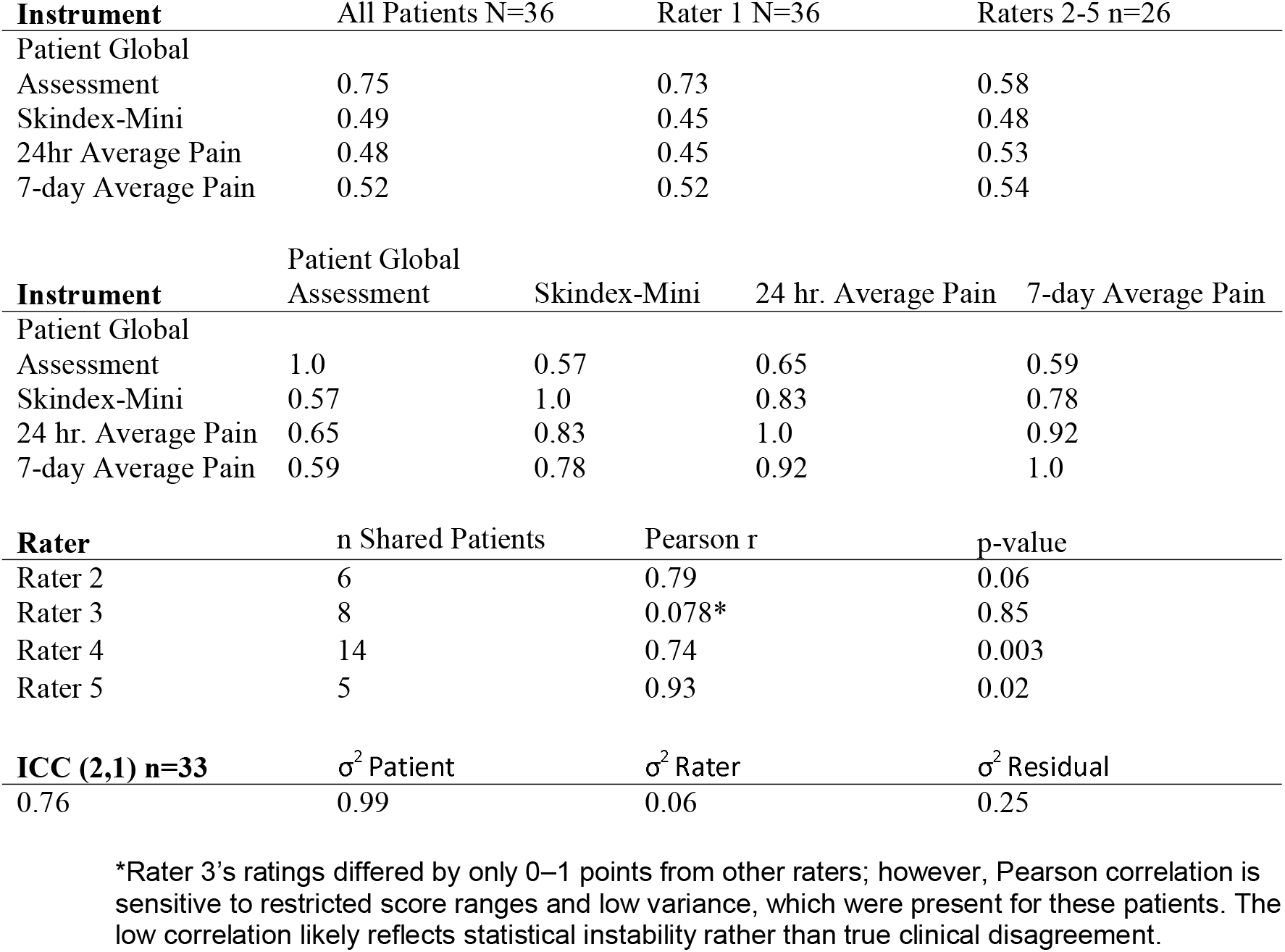
Construct validity and inter-rater reliability of the IGAPg morphological tool. Correlations between IGAPg scores and patient-reported outcomes (PGA, Skindex-Mini, and average pain) demonstrate moderate-to-strong construct validity. Values are shown for all raters, Rater 1 alone, and Raters 2-5. Inter-item PRO correlations confirm internal consistency. Pearson correlations between Rater 1 and Raters 2-5 are reported with shared patient counts and p-values (p < 0.05 was considered significant). ICC(2,1) indicated good inter-rater reliability with most score variability attributable to differences between patients with minimal variability due to rater differences and a moderate residual component. Inter-item patient reported outcomes correlations confirm difference in measured constructs.

We report the development and initial validation of a novel IGA for PG, the **IGAPg**© (available https://data.mendeley.com/datasets/3bbm5cgtgh/1). An international multistakeholder panel guided its creation. A core team (MEJ, AM, AOL) designed the instrument based on prior research into key clinical signs of PG severity, with critical feedback and input provided by a review team (coauthors).^3-5^ While core outcome sets are in development, existing PG assessments remain limited to ulcer size or non–PG-specific measures, falling short of FDA expectations and underscoring the need for a validated PG-specific tool. The **IGAPg**© incorporates objective ulcer features—depth, drainage, discoloration, and undermining—with location and extent used to resolve indeterminate cases. Standardized rater training materials were also developed.

Initial validation assessed construct validity and reliability. The **IGAPg**© was evaluated in 36 patients by an expert clinician (AOL), with patients completing the Patient Global Assessment (PGA), Skindex-Mini, and numeric rating scales (NRS) for 24-hour and 7-day average pain. Pearson correlations were used to assess construct validity (supplementary methods).

A subset of 26 patients was independently evaluated by the expert clinical rater (AOL) and four additional raters (Fig 1). All assessments were conducted during routine clinic visits with raters concurrently present; evaluations were based on direct clinical examination rather than photographs. Inter-rater reliability was examined using a two-way random-effects intraclass correlation coefficient (ICC [2,1]), implemented within a linear mixed-effects model that included patient ID and rater as random intercepts. This approach appropriately accommodates the unbalanced design, in which not all raters evaluated every patient. Analyses were conducted using Python (v3.11).

**Figure 1.**
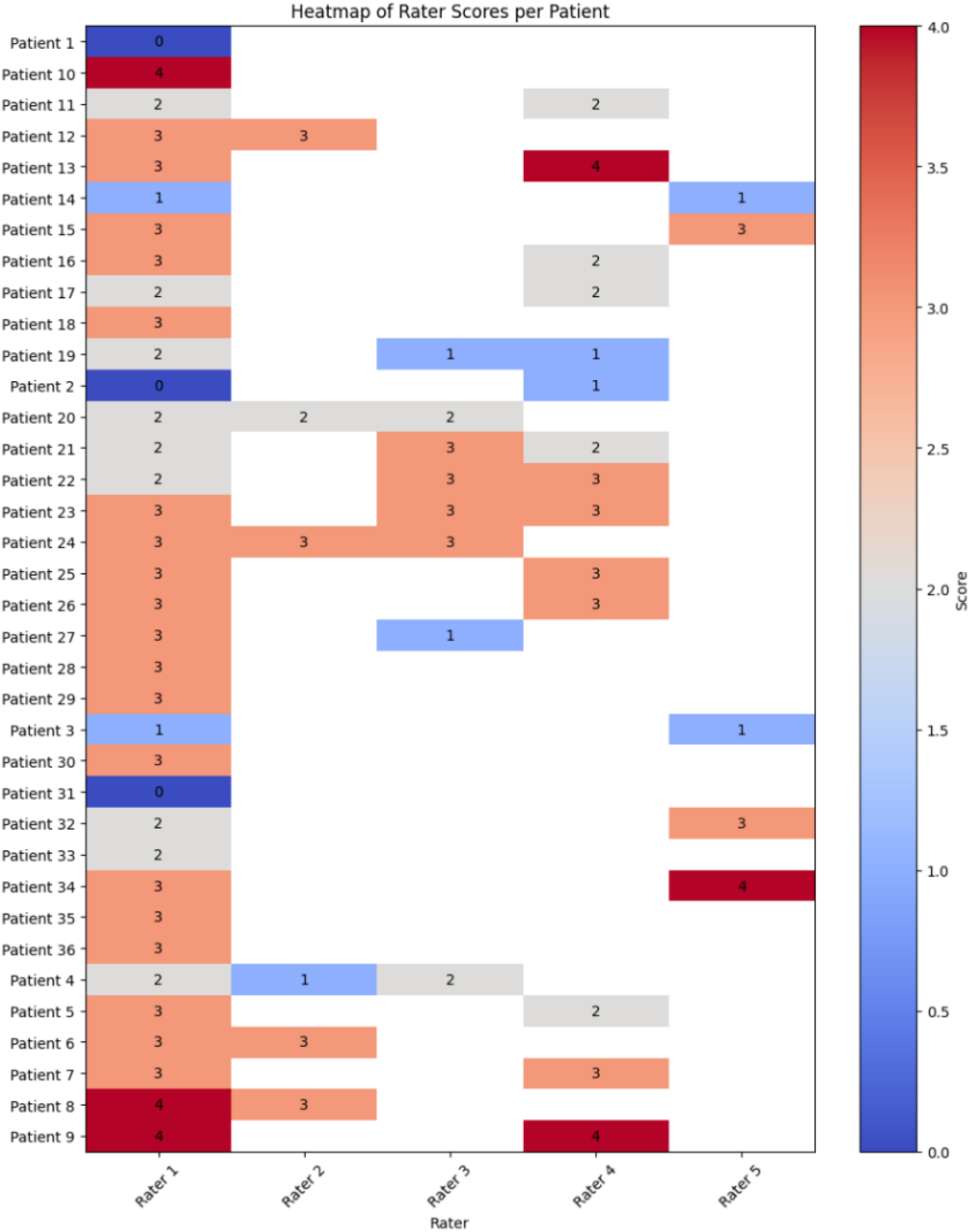
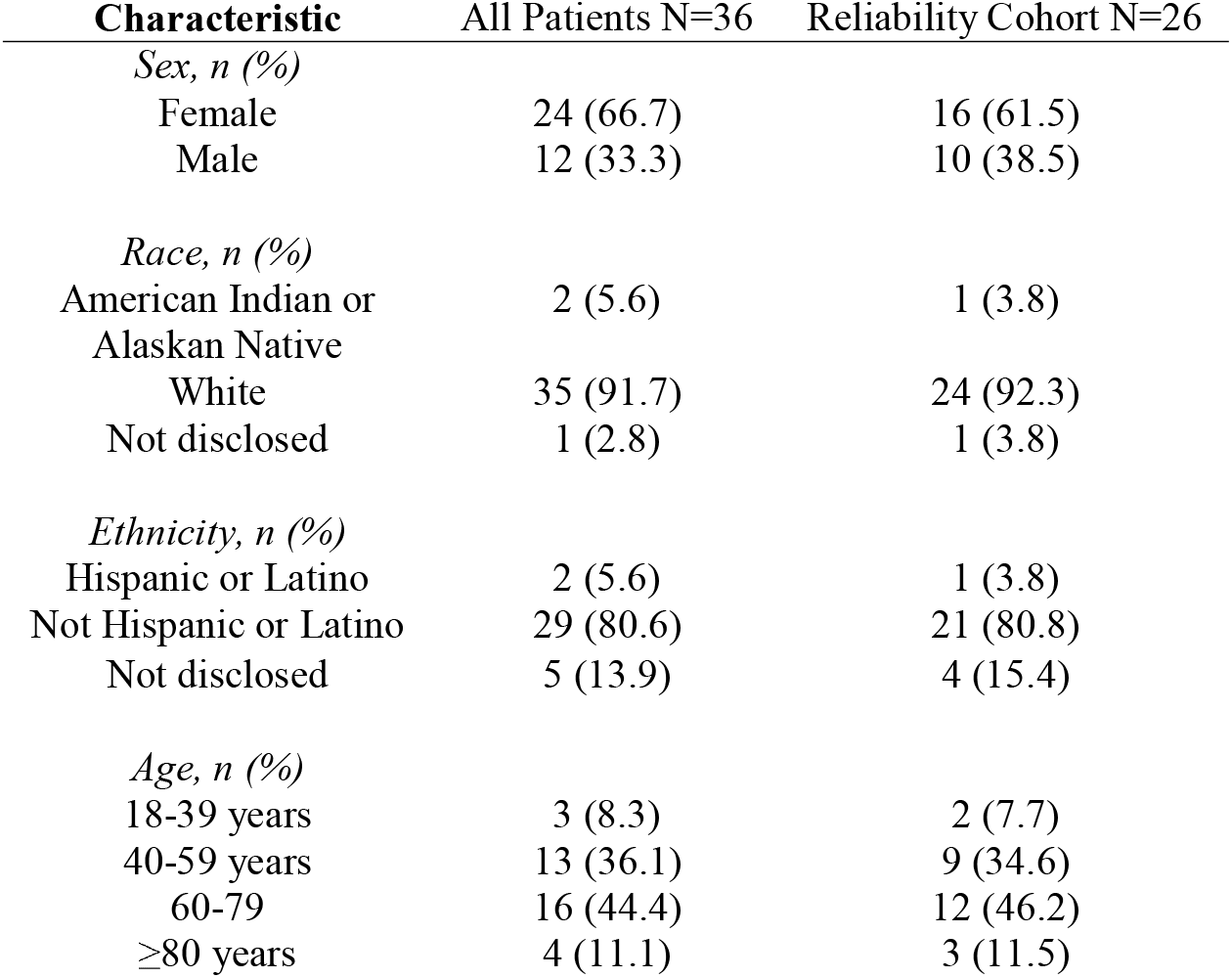
**Top**-Heatmap of clinical rater IGAPg scores and rater characteristics. Warmer colors indicate higher severity scores and cooler colors indicate lower scores. Rater 1: Dermatologist instrument creator; Raters 2, 4, 5: Dermatology trainees; Rater 3: Attending dermatologist. **Bottom-**Pyoderma Gangrenosum patient participant demographics. The reliability cohort includes patients assessed by two or more raters.

**IGAPg**© scores strongly correlated with the PGA when assessed by a PG expert dermatologist (Pearson’s r = 0.73; Fig 1). When averaged across all raters, the correlation remained strong (r = 0.69; Fig 1). Moderate correlations were observed between IGAPg and Skindex-Mini (r = 0.49), 24-hour pain NRS (r = 0.48), and 7-day pain NRS (r = 0.52), reflecting the difference in constructs being measured (Fig 1). Inter-rater reliability was high, with an ICC of 0.76 (Fig 1). Raters reported that the measure was comprehensive, comprehensible, and quick to use. Reliability and construct validity data are presented in Table 1. The IGAPg© was developed for dermatologists and trainees and requires familiarity with PG-specific ulcer morphology; intentionally designed for use in clinical research and trials.

Despite limitations related to modest sample size and rater homogeneity in training background, the **IGAPg**© demonstrated strong construct validity and reliability. Future directions include refining training materials and assessment content, as well as conducting validation studies that incorporate structured patient-investigator engagement. Overall, the **IGAPg**© represents a promising tool for assessing PG severity and treatment response.

## Supporting information

IGAPg tool, image atlas and examples

## Data Availability

All data produced in the present study are available upon reasonable request to the authors

