## Supplementary material for "Initial development and pragmatic clinical validation of a static disease severity instrument for pyoderma gangrenosum: Investigator Global Assessment for PG (IGAPg)": IGAPg tool, image atlas and examples

The score is selected using the descriptors below that best describe the overall appearance of the majority of the involved area at a given time point. For a rating of clear, all three morphological descriptions must apply to all of the involved tissue. It is not necessary that all characteristics under *Morphological Description* be present. Please include all affected areas in their totality.

| Score | Morphological Description |
|---|---|
| 0-Clear | Complete re-epithelization.<br>No redness.<br>No accumulation of drainage on the surface of the ulcer. |
| 1-Almost Clear | Erosion.<br>Light redness around the ulcer borders.<br>The wound surface is moist. |
| 2-Mild | Ulceration extends to the dermis.<br>Dull redness around the ulcer borders.<br>Presence of pooling regions of exudate on the surface of the ulcer. |
| 3-Moderate | Ulceration extends to the subcutaneous tissue.<br>Bright redness around ulcer borders.<br>Presence of a confluent layer of exudate covering the wound surface.<br>Undermining may be present in <50% of the ulcer borders. |
| 4-Severe | Presence of muscle, bone, or tendon in the ulcer.<br>Violaceous/gunmetal grey discoloration around ulcer borders. Blistering may be present in the periwound area.<br>Evidence of a layer of exudate extending beyond wound borders.<br>Undermining may be present in up to 100% of the wound borders. |

In indeterminate cases, please use extent (size and/or quantity of lesions) and/or involvement of special sites (head, neck, and/or genitalia) to differentiate between scores

**© 2025, Oregon Health & Science University**

### IGAPg

The score is selected using the descriptors below that best describe the overall appearance of the majority of the involved area at a given time point. For a rating of clear, all three morphological descriptions must apply to all of the involved tissue. It is not necessary that all characteristics under *Morphological Description* be present. Please include all affected areas in their totality.

| Score | Morphological Description |
|---|---|
| 0-Clear | Complete re-epithelization.<br>No redness.<br>No accumulation of drainage on the surface of the ulcer. |
| 1-Almost Clear | Erosion.<br>Light redness around the ulcer borders.<br>The wound surface is moist. |
| 2-Mild | Ulceration extends to the dermis.<br>Dull redness around the ulcer borders.<br>Presence of pooling regions of exudate on the surface of the ulcer. |
| 3-Moderate | Ulceration extends to the subcutaneous tissue.<br>Bright redness around ulcer borders.<br>Presence of a confluent layer of exudate covering the wound surface.<br>Undermining may be present in <50% of the ulcer borders. |
| 4-Severe | Presence of muscle, bone, or tendon in the ulcer.<br>Violaceous/gunmetal grey discoloration around ulcer borders. Blistering may be present in the periwound area.<br>Evidence of a layer of exudate extending beyond wound borders.<br>Undermining may be present in up to 100% of the wound borders. |

In indeterminate cases, please use extent (size and/or quantity of lesions) and/or involvement of special sites (head, neck, and/or genitalia) to differentiate between scores

© 2025, Oregon Health & Science University

**IGAPg lesion severity atlas**

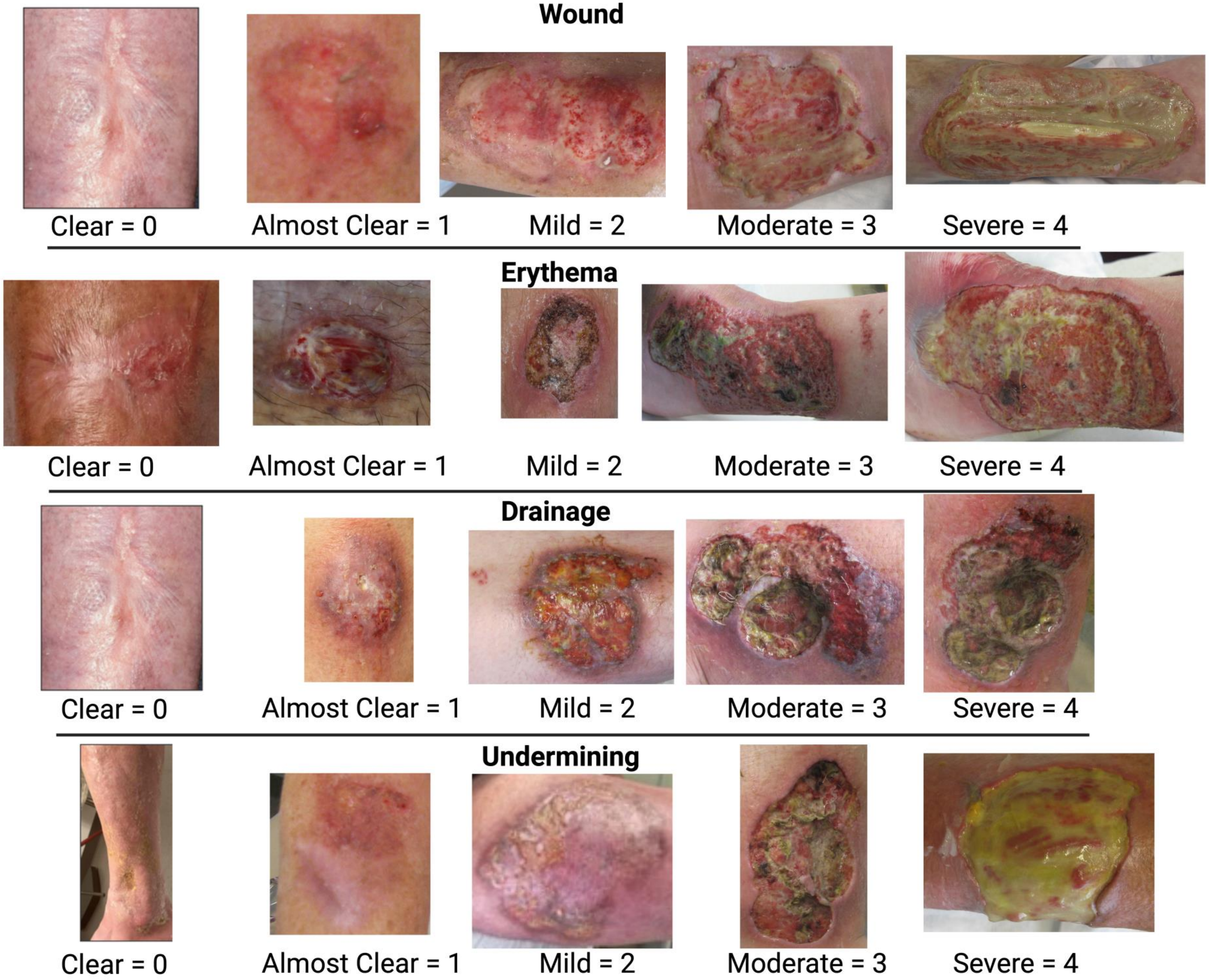

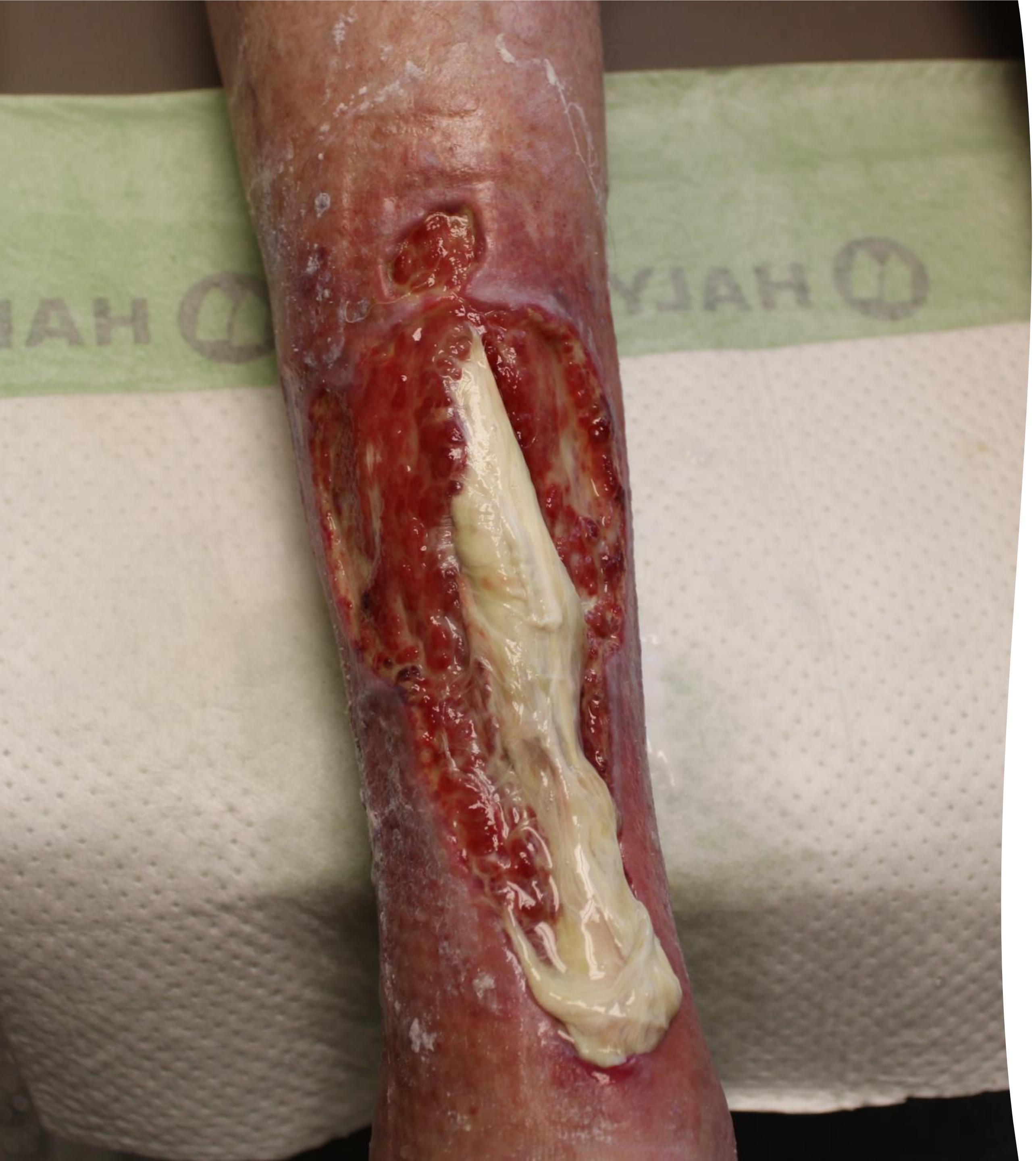

- Wound: 4
- Erythema: 3
- Drainage: 3
- Undermining: 3

- **IGAPg score: 4**

**SEVERE**

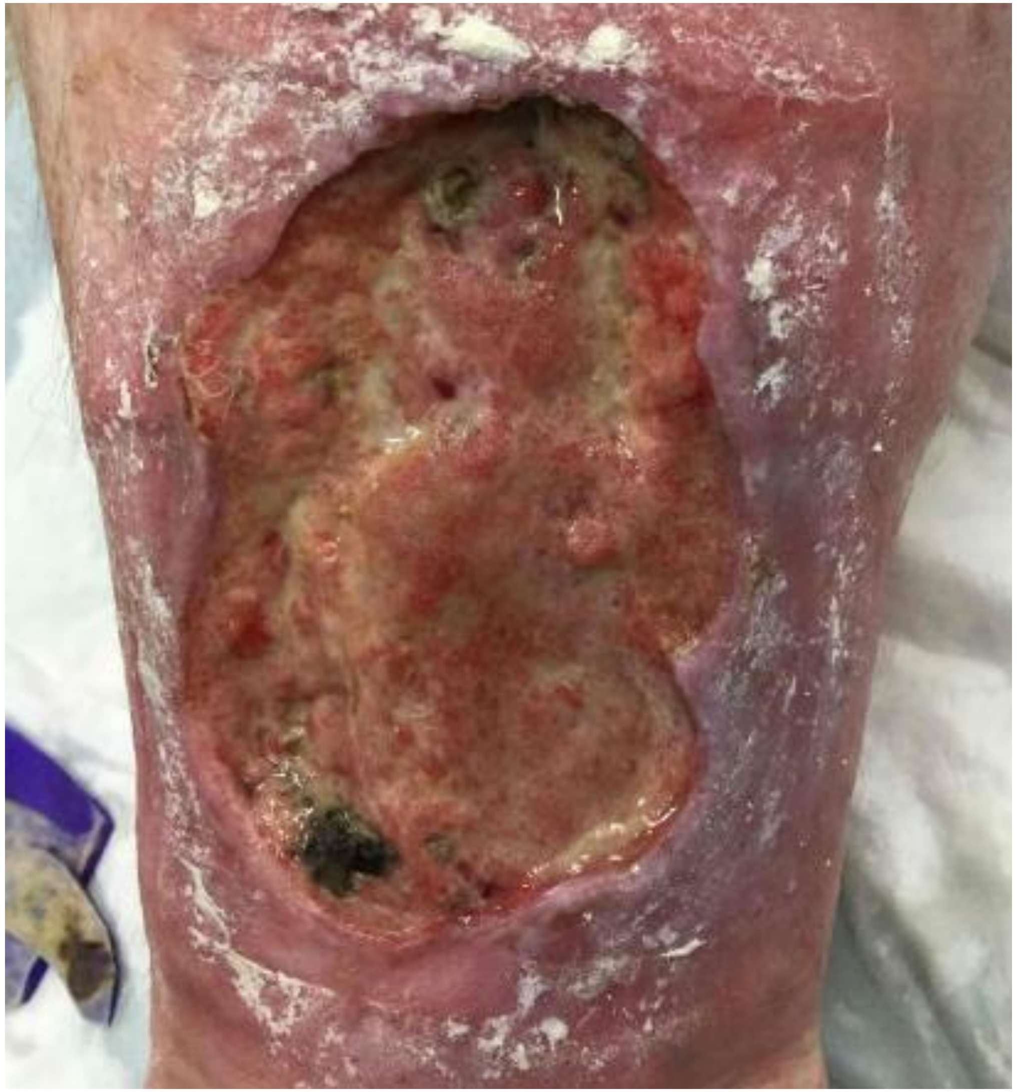

- Wound: 3
- Erythema: 3
- Drainage: 2
- Undermining: 3

- **IGAPg score: 3**

**MODERATE**

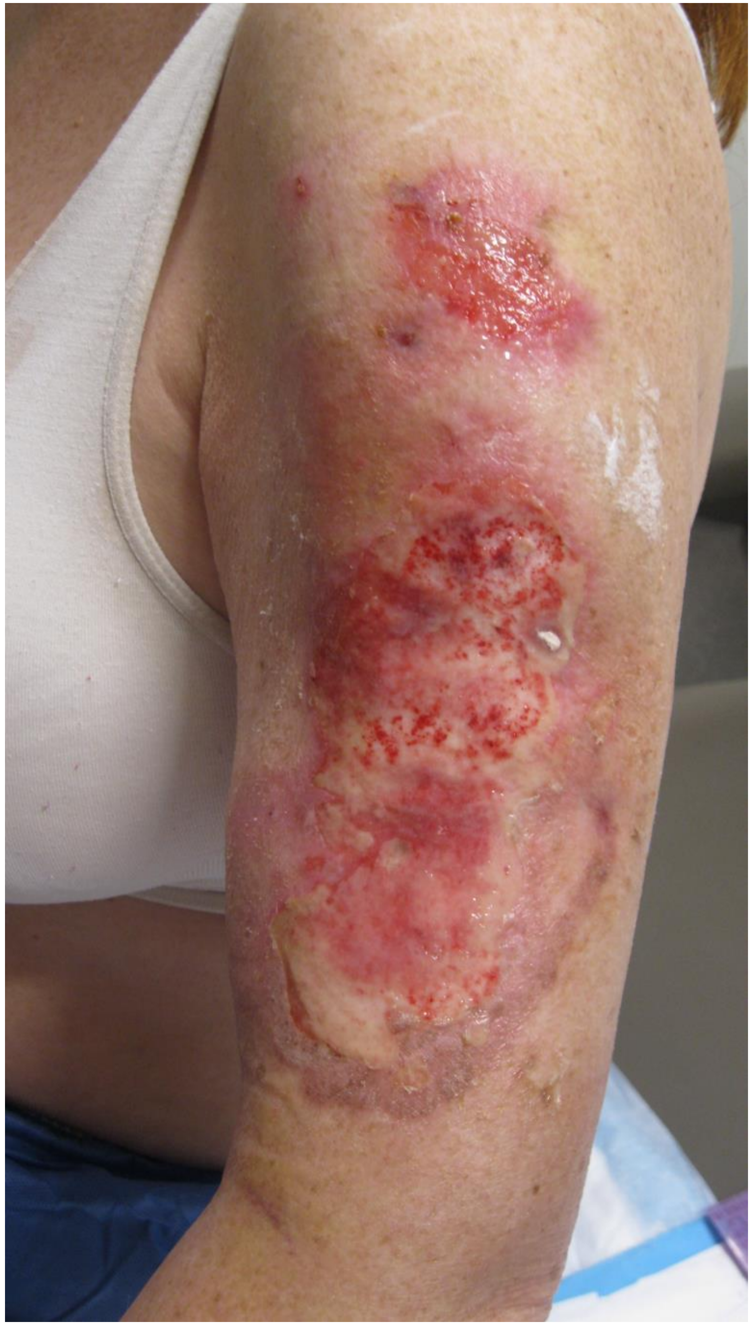

- Wound: 2
- Erythema: 2
- Drainage: 3
- Undermining: 0
- **IGAPg score: 2**

**MILD**

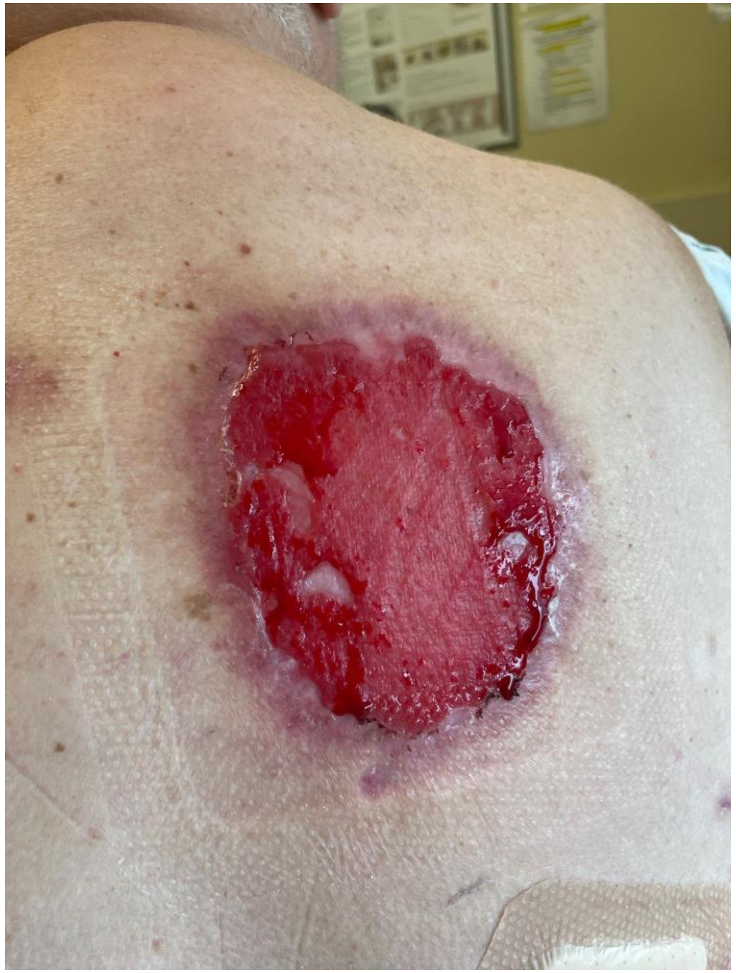

- Wound: 2
- Erythema: 1
- Drainage: 1
- Undermining: 0

- **IGAPg score: 1**

**ALMOST CLEAR**

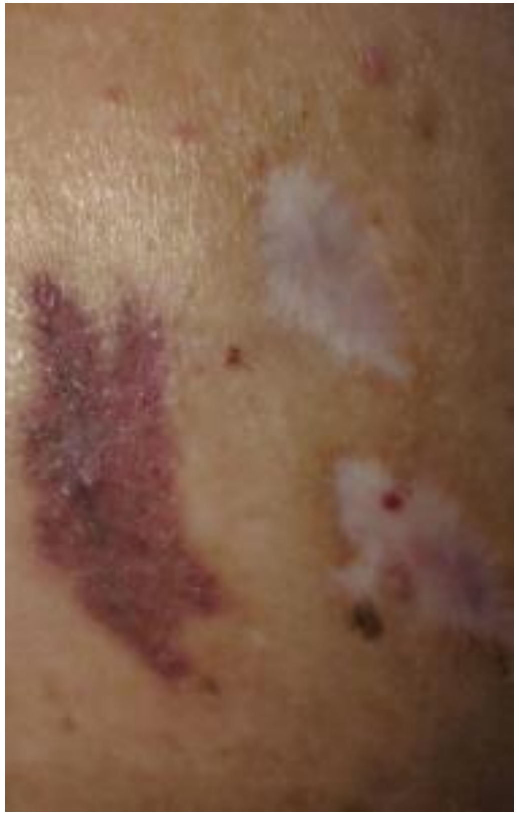

- Wound: 0
- Erythema: 0
- Drainage: 0
- Undermining: 0

- **IGAPg score: 0**

**CLEAR**
